# Lower insulin resistance in Chinese patients with severe major depressive disorder: associations with the inflammatory response

**DOI:** 10.1101/2025.10.10.25337709

**Authors:** Yueyang Luo, Mengqi Niu, Tangcong Chen, Jing Li, Abbas F. Almulla, Yingqian Zhang, Michael Maes

## Abstract

**Background:** Major depressive disorder (MDD) is widely acknowledged as stemming from the dysregulation of neuroimmune, metabolic, and oxidative stress (NIMETOX) pathways. The objective of this study was to examine insulin metabolism in Chinese patients with MDD and to clarify the relationship between insulin resistance and the acute phase protein (APP) response, as shown by the negative APPs albumin and transferrin.

**Methods:** This investigation utilized a cross-sectional case-control approach, enrolling 125 inpatients with MDD and 40 healthy controls.

**Results:** Patients with MDD exhibited markedly reduced levels of fasting plasma glucose, insulin, and insulin resistance, and heightened insulin sensitivity, in comparison to healthy controls. The significance of these alterations persisted after controlling for metabolic syndrome, body mass index (BMI), and age, but was nullified with adjustment for both negative APPs. We determined that 41.4% of the variance in insulin resistance was accounted for by elevated levels of BMI, albumin, transferrin, and age. Insulin resistance was significantly and inversely associated with weight loss. We found that 27.7% of the variance in overall depression severity was accounted for by adverse childhood experiences (positive correlation) and insulin resistance (negative correlation).

**Conclusions:** This work demonstrates that Chinese MDD patients display increased insulin sensitivity and reduced insulin resistance, with these alterations being associated with a modest smoldering inflammatory response. MDD is characterized by a hormetic response that enhances insulin efficacy, hence optimizing glucose consumption to sustain normal organ function. It is incorrect to claim that MDD is intrinsically associated with increased insulin resistance.

## Introduction

Major depressive disorder (MDD) is increasingly recognized as a result of dysregulation of neuroimmune, metabolic, and oxidative stress (NIMETOX) pathways (Maes, Almulla, et al., 2025a; Maes, Jirakran, et al., 2025b). Current research suggests that MDD is closely associated with chronic low-grade inflammation, insulin resistance (IR), and multiple cardiometabolic diseases, including metabolic syndrome (MetS), type 2 diabetes mellitus (T2DM), and cardiovascular disease (Ma et al., 2025; Maes, Almulla, et al., 2025a; Maes, Niu, et al., 2025c). Within this immunometabolic framework, activation of M1 macrophages and elevated levels of proinflammatory cytokines, such as interleukin (IL)-6, IL-1β, and tumor necrosis factor (TNF)-α, might impair insulin signaling, triggering peripheral and central insulin resistance that leads to hyperglycemia and compensatory hyperinsulinemia (Maes, Almulla, et al., 2025a; Maes, Jirakran, et al., 2025b). This inflammatory response triggers an acute-phase reaction in the liver, reducing the levels of negative acute-phase proteins (APPs), particularly albumin and transferrin (Maes, 1993).

Although scientific research often suggests a bidirectional association between insulin resistance and MDD, the findings vary across studies (Fanelli et al., 2025). Some reviews indicate significantly elevated fasting insulin levels and HOMA-IR indices in MDD patients (Fernandes et al., 2022), whereas other studies did not find increased insulin resistance indices in MDD compared to controls (de Melo et al., 2017; Maes, Jirakran, et al., 2025b; Morelli et al., 2021). A meta-analysis showed that during the acute phase of depression, patients had significantly elevated insulin levels, HOMA-IR indices, and insulin variability (Fernandes et al., 2022). In contrast, these markers showed no significant changes during remission or following antidepressant treatment (Fernandes et al., 2022). However, the latter analysis has a key methodological limitation: the strategy for controlling confounding factors relied solely on matching for sex, age, and BMI during the study design phase, without further adjusting for MetS, overweight or obesity, in the statistical models. The inclusion of people with MetS or high BMI values in the analyses may have caused bias in metabolic data resulting in biased findings (Maes et al., 2024; Maes, Jirakran, et al., 2025b).

Nevertheless, the associations between insulin resistance and MDD go far beyond the possible differences between MDD and healthy controls. Thus, increased insulin resistance in MDD contributes to increased oxidative and nitrosative stress, and together with enhanced activation of cytokine networks, leads to increased severity of the affective and physio-somatic symptoms in MDD (Maes, Jirakran, et al., 2025b; Morelli et al., 2021). Such changes may be explained by intertwined causal relationships between insulin resistance, oxidative stress and activated cytokine networks (Maes, Jirakran, et al., 2025b).

The inconsistent association between insulin resistance and MDD limits its reliability as a robust biomarker of MDD. In contrast, the negative APP response with low albumin and transferrin levels demonstrate great consistency in their association with MDD (Maes et al., 1991; Wainberg et al., 2021). These two negative APPs are closely linked to immune activation, adverse childhood experiences (ACEs), and the severity of somatic and affective symptoms (Almulla et al., 2025; Maes, 1993). While alterations in negative APPs (albumin and transferrin) have consistently been linked to MDD and immune activation, their interactions with insulin resistance in MDD patients remain unclear. In addition, research specifically examining the relationship between insulin resistance and inflammation in Chinese MDD patients remains scarce.

Thus, this study aimed to investigate the insulin resistance status in Chinese MDD patients and further elucidate the association between insulin resistance and the negative APP response, as reflected by the negative APPs albumin and transferrin. Our hypothesis is that changes in insulin resistance are associated with the severity of MDD and that these changes are associated with the negative APP response.

## Methods

### Participants

This study employed a cross-sectional case–control design and enrolled 165 participants, including 125 individuals diagnosed with major depressive disorder (MDD) and 40 healthy controls. Recruitment was conducted at the Psychiatric Center of Sichuan Provincial People’s Hospital, Chengdu, China. The inclusion criteria for MDD participants were as follows: (1) a confirmed diagnosis of MDD according to the Diagnostic and Statistical Manual of Mental Disorders, Fifth Edition (DSM-5); (2) a score greater than 18 on the 21-item Hamilton Depression Rating Scale (HAMD-21) (Hamilton, 1960); (3) age between 18 and 65 years, regardless of sex; and (4) the ability to provide written informed consent, with guardian consent when required. Healthy controls were matched to MDD patients based on sex, age, body mass index (BMI), and education level, and were recruited from among hospital staff and their acquaintances.

The exclusion criteria for both MDD patients and healthy controls included: (1) pregnancy or lactation; (2) a history of acute infections or recent surgeries within the last three months; (3) severe allergic reactions in the preceding month; (4) treatment with immunosuppressive or immunomodulatory agents, including glucocorticoids; (5) use of therapeutic-dose antioxidants or omega-3 supplements in the last three months; (6) frequent or chronic analgesic use; (7) serious medical illnesses such as autoimmune disorders, systemic lupus erythematosus, rheumatoid arthritis, type 1 diabetes, inflammatory bowel disease, psoriasis, chronic obstructive pulmonary disease, or malignancy; (8) neurological disorders including epilepsy, stroke, brain tumors, Parkinson’s disease, Alzheimer’s disease, or multiple sclerosis; (9) personality or developmental disorders (e.g., borderline, antisocial, or severe intellectual disability); (10) other major psychiatric diagnoses, such as bipolar disorder, schizophrenia, schizoaffective disorder, psycho-organic conditions, or substance-use disorders (except nicotine dependence). Furthermore, healthy controls were excluded if they had a history of MDD or dysthymia, any DSM-IV anxiety disorder, or a family history of mood disorders, suicide, or substance use disorders other than nicotine dependence.

Ethical approval for this study was obtained from the Institutional Review Board of Sichuan Provincial People’s Hospital, Chengdu, China [Ethics (Research) 2024-203] and all participants provided written informed consent per the Declaration of Helsinki.

### Clinical Assessments

All participants underwent a semi-structured interview conducted by a trained physician to collect demographic and clinical information, including age, sex, education, medical and psychiatric history, and family history. Psychiatric diagnoses were assessed using the Mini International Neuropsychiatric Interview (MINI) (Sheehan et al., 1998), which screens for both current and lifetime psychiatric disorders according to the DSM-IV and ICD-10 criteria.

On the same day, a single evaluator assessed depression, anxiety, and somatic symptoms. Depression severity was measured using the total score on the 21-item Hamilton Depression Rating Scale (HAMD) (Hamilton, 1960), and anxiety severity was assessed using the Hamilton Anxiety Rating Scale (HAMA) (Hamilton, 1959). Self-reported anxiety was evaluated using the state version of the State-Trait Anxiety Inventory (STAI) (Spielberger et al., 1971). Fibromyalgia and Chronic Fatigue Syndrome (CFS) symptoms were quantified using the 12-item Fibro Fatigue Scale (FF) (Zachrisson et al., 2002).

To minimize the influence of overlapping somatic items across scales, *pure scores* were derived for each affective domain. Pure HAMD and pure HAMA were computed by selecting items specific to depression, anxiety, and subjective anxiety, while excluding confounding somatic items. Pure HAMD was computed as the sum of the individual items related to depression, guilt, suicidal ideation, and loss of interest, excluding any somatic items. Pure HAMA score is the sum of items that assess anxiety, tension, fear, and anxious behavior. Pure FF score is calculated by summing the z-scores of items related to muscle pain, muscle tension, flu-like symptoms, fatigue, headache, autonomic and gastro-intestinal symptoms. To create an integrated clinical severity index, we computed a z unit-based composite score by summing the z-scores of the pure HAMD, pure HAMA, pure STAI, and pure FF (labeled as overall severity of depression or OSOD. Weight loss was assessed using the relevant items from the HAMD. Adverse childhood experiences (ACEs) were evaluated using the Childhood Trauma Questionnaire-Short Form (CTQ-SF) (Bernstein et al., 2003). The total ACEs score was computed by summing the five subscales: emotional abuse, physical abuse, sexual abuse, emotional neglect, and physical neglect (Vasupanrajit et al., 2024).

Physical measurements, including height, weight, and blood pressure were recorded. BMI was calculated by dividing the weight in kilograms by the height in meters squared. Waist circumference (WC) was measured at the midpoint between the iliac crest and lowest rib. An integrated measure of body composition was obtained by calculating a composite z-score combining BMI and WC (z BMI + z WC). MetS was defined based on the 2009 Joint Scientific Statement of the American Heart Association and the National Heart, Lung, and Blood Institute (Alberti et al., 2009), requiring at least three of the following criteria: (1) triglycerides ≥ 150 mg/dL; (2) WC ≥ 90 cm (men) or ≥ 80 cm (women); (3) elevated blood pressure (systolic > 130 mmHg, diastolic > 85 mmHg, or current antihypertensive medication use); (4) HDL cholesterol < 40 mg/dL (men) or < 50 mg/dL (women); and (5) fasting glucose ≥ 100 mg/dL or a diagnosis of diabetes. Participants were ranked based on the number of MetS criteria they fulfilled.

#### Assays

Fasting (10 hours) venous blood (30 mL) was collected from each participant between 6:30 and 8:00 a.m. using disposable syringes and serum tubes. Following centrifugation at 3,500 rpm, the serum was aliquoted into Eppendorf tubes and stored at −80[°C until analysis. Fasting blood glucose (FBG) was measured using the glucose assay kit (glucose oxidase method, Gcell, Beijing Strong Biotechnologies, Inc.) on a fully automated biochemical analyzer (ADVIA 2400, Siemens Healthcare Diagnostics Inc.) with the intra-assay and inter-assay analytical coefficients of variation (CVs) of 3.2% and 6.8% respectively. Serum insulin was assayed using Atellica IM insulin assay kit (direct chemiluminescence method, Siemens Healthcare Diagnostics Inc.) on Atellica IM fully automated chemiluminescence immunoassay analyzer (Siemens Healthcare Diagnostics Inc.). The intra-assay and inter-assay analytical CVs are 1.8% and 3.6% respectively. Serum triglycerides (TG) were measured by TG assay kits (GPO-PAP method, Gcell, Beijing Strong Biotechnologies, Inc.) on a fully automated biochemical analyzer (ADVIA 2400, Siemens Healthcare Diagnostics Inc.) with the intra-assay and inter-assay analytical coefficients of variation (CVs) of 3.0% and 7.0% respectively.

Basal insulin resistance was assessed using HOMA2-IR the Homeostasis Model Assessment of Insulin Resistance), with beta-cell function determined by HOMA2-β and insulin sensitivity by HOMA2-IS. Calculations were performed using the HOMA2 calculator (http://www.dtu.ox.ac.uk). Using z unit-based composite scores we computed indices of insulin resistance as z FBG + z insulin and insulin sensitivity as z insulin – z FPG (Maes, Jirakran, et al., 2025b). The TyG index, another index assessing insulin resistance, was determined using the formula: ln{fasting triglyceride (mg/dL)×fasting plasma glucose (mg/dL)/2}(Abbasi & Reaven, 2011). Additionally, the Quantitative Insulin Sensitivity Check Index (QUICKI) was employed as a validated method for evaluating insulin sensitivity in both diabetic and non-diabetic individuals, including those with obesity (Muniyappa et al., 2008). The QUICKI score is calculated using the formula: QUICKI = 1 / {log (Fasting Insulin) + log (Fasting Glucose)}. This index demonstrates a strong correlation with glucose clamp studies (r = 0.78) (Muniyappa et al., 2008).

Serum transferrin was measured using an immunoturbidimetric assay (DIAYS DIAGNOSTIC SYSTEM, SHANGHAI CO., LTD) on a fully automated biochemical analyzer (ADVIA 2400, Siemens Healthcare Diagnostics Inc.) with a sensitivity of 0.03 g/L. The intra-assay and inter-assay analytical coefficients of variation (CVs) were 1.96% and 0.67%, respectively. Serum albumin was measured using the Bromocresol Green Method kit (Beijing Strong Biotechnologies, Inc.) on a fully automated biochemical analyzer (ADVIA 2400, Siemens Healthcare Diagnostics Inc.) with the intra-assay and inter-assay CVs of 1.20% and 2.10%, respectively. To compute an integrated index of the negative APPs response, a composite z-score was calculated by adding the z-scores for albumin and transferrin.

#### Statistics

Pearson’s correlation calculation was employed to evaluate the connections between continuous variables. Contingency tables were used to analyze the associations between categorical variables. Differences in continuous variables among groups were assessed using analysis of variance (ANOVA). False Discovery Rate (FDR) was applied to correct multiple comparisons. The study considered potential confounders such as age, sex, smoking status, and BMI. Binary logistic regression analysis was employed to predict MDD (controls as reference group) using insulin biomarkers, albumin, transferrin, ACEs, and demographic data. Key outcomes were Nagelkerke pseudo-R² (used as effect size), Wald statistics with corresponding p-values, odds ratios (OR) with 95% confidence intervals (CI), and unstandardized regression coefficients (B) with their standard errors (SE). Multiple regression analysis was used to predict the insulin biomarkers with albumin, transferrin, weight loss, and metabolic data as explanatory variables. Another series of multiple regression analysis examined the effects of insulin biomarkers and other predictors on the clinical rating scale scores. Both manual and automatic stepwise methods were employed. The automated procedure involved linear modeling with measures to prevent overfitting, with entry and removal criteria set at p = 0.05 and p = 0.07, respectively. The model outputs included standardized beta coefficients, degrees of freedom (df), p-values, R², and F-statistics. Heteroskedasticity was evaluated using the White and modified Breusch-Pagan tests, while collinearity was assessed using the tolerance and variance inflation factor (VIF). All analyses were two-tailed with a significance level of 0.05 for all tests. IBM SPSS Statistics 30 (Windows) was used for all analyses, and data transformations, including log10, square root, rank-order, and Winsorization, were applied where needed.

## Results

### Socio-demographic and Clinical Characteristics

**Table 1** shows the socio-demographic and clinical characteristics of the study participants. No significant differences were observed between MDD and HC in terms of age, gender distribution, the prevalence of MetS, MetS ranking, BMI, WC, the z BMI + z WC score, history of smoking, or years of education. However, the MDD group demonstrated a significantly higher weight loss score compared to the HC group. Clinical assessments revealed significantly higher total scores on HAMD, HAMA, FF, and STAI in MDD patients relative to controls. Furthermore, MDD patients reported significantly higher levels of ACEs and a greater OSOD score. Serum albumin and transferrin levels were markedly lower in the MDD group, accompanied by a significant reduction in the z albumin + z transferrin score.

**Table 1.** Demographic and clinical profile of patients with major depressive disorder (MDD) and healthy controls (HC).

| Variables | HC (n=40) | MDD (n=125) | F / $\chi^2$ | df | p-value |
| --- | --- | --- | --- | --- | --- |
| Age (years) | 37.1 (13.7) | 35.7 (12.1) | 0.37 | 1/163 | 0.542 |
| Gender (m/f) | 13/27 | 38/87 | 0.06a | 1 | 0.845 |
| Metabolic syndrome (MetS) (No/Yes) | 29/10 | 102/22 | 1.173a | 1 | 0.355 |
| Ranking MetS | 1.59 (1.46) | 1.42 (1.26) | 0.50 | 1/161 | 0.479 |
| Body Mass Index (BMI) (kg/m<sup>2</sup>) | 23.52 (4.07) | 22.299 (3.36) | 3.58 | 1/163 | 0.060 |
| Waist circumference (WC) (cm) | 79.38 (11.73) | 78.27 (11.32) | 0.28 | 1/163 | 0.595 |
| z BMI + z WC (z score) | 0.332 (2.020) | -0.106 (1.840) | 1.64 | 1/163 | 0.202 |
| Weight loss (z score) | -0.605 (0.000) | 0.194 (1.080) | 21.80 | 1/163 | <0.001 |
| History of smoking (No/Yes) | 37/3 | 101/24 | 3.03a | 1 | 0.091 |
| Education years | 13.9 (4.3) | 13.5 (3.3) | 0.26 | 1/163 | 0.613 |
| Albumin (g/L) | 43.64 (1.99) | 41.18 (3.05) | 22.44 | 1/161 | <0.001 |
| Transferrin (mg/dL) | 258.76 (37.44) | 228.25 (38.66) | 18.79 | 1/162 | <0.001 |
| z albumin + z transferrin (z score) | 0.723 (0.805) | -0.227 (0.948) | 31.92 | 1/161 | <0.001 |
| Triglycerides (mmol/L) | 1.39 (0.10) | 1.36 (0.06) | 0.205 | 1/161 | 0.479 |
| Total HAMD | 0.9 (2.0) | 28.5 (5.2) | MWUT | - | <0.001 |
| Pure HAMD | 0.0 (0.0) | 8.26 (4.04) | MWUT | - | <0.001 |
| Total HAMA | 1.1 (3.0) | 26.6 (8.6) | MWUT | - | <0.001 |
| Pure HAMA | 0.1 (0.5) | 9.0 (3.2) | MWUT | - | <0.001 |
| Total FF | 1.4 (2.9) | 29.9 (9.9) | MWUT | - | <0.001 |
| Pure FF | 0.5 (1.0) | 11.0 (6.0) | MWUT | - | <0.001 |
| STAI state | 36.8 (8.3) | 58.1 (10.4) | 137.23 | 1/153 | <0.001 |
| Total ACEs | -0.746 (0.821) | 0.239 (0.935) | 35.55 | 1/163 | <0.001 |
| OSOD (PC score) | -1.518 (0.237) | 0.528 (0.491) | 640.21 | 1/153 | <0.001 |

Data are presented as mean (SD); F: F-statistic from ANOVA; $\chi^2$ : Chi-square test statistic; MWUT: Mann–Whitney U test statistic.
ACE: adverse childhood experiences, HAMD: Hamilton Depression Rating Scale, HAMA: Hamilton Anxiety Rating Scale, FF: Fibro-Fatigue, STAI: State-Trait Anxiety Inventory, OSOD (PC score): overall severity of depression (principal component score).

### Insulin biomarkers in MDD

**Table 2** presents the results of GLM analyses examining the associations between metabolic indicators and MDD, both before and after adjustment for MetS, BMI, age, transferrin, and albumin. Prior to adjustment, MDD patients showed significantly lower levels of FPG, insulin, and z FPG + z INS and HOMA2-IR scores, alongside significantly higher levels of insulin sensitivity as indicated by HOMA2-IS and QUICKI, compared to healthy controls. All these differences remained significant after FDR p-correction (all at p< 0.0135). After controlling for MetS, BMI, and age all those differences remained significant, whilst also HOMA-2β became significant (decreased in MDD). After excluding subjects with MetS, the decreases in insulin resistance and increased in insulin sensitivity (p<0.01) in MDD versus controls remained significant. The differences also remained significant after FDR p=correction (all at p=0.0167). After adding the negative acute phase proteins as covariates only HOMA2-IR remained significant. Nevertheless, after FDR p correction, this variable was no longer significant.

**Table 2.**
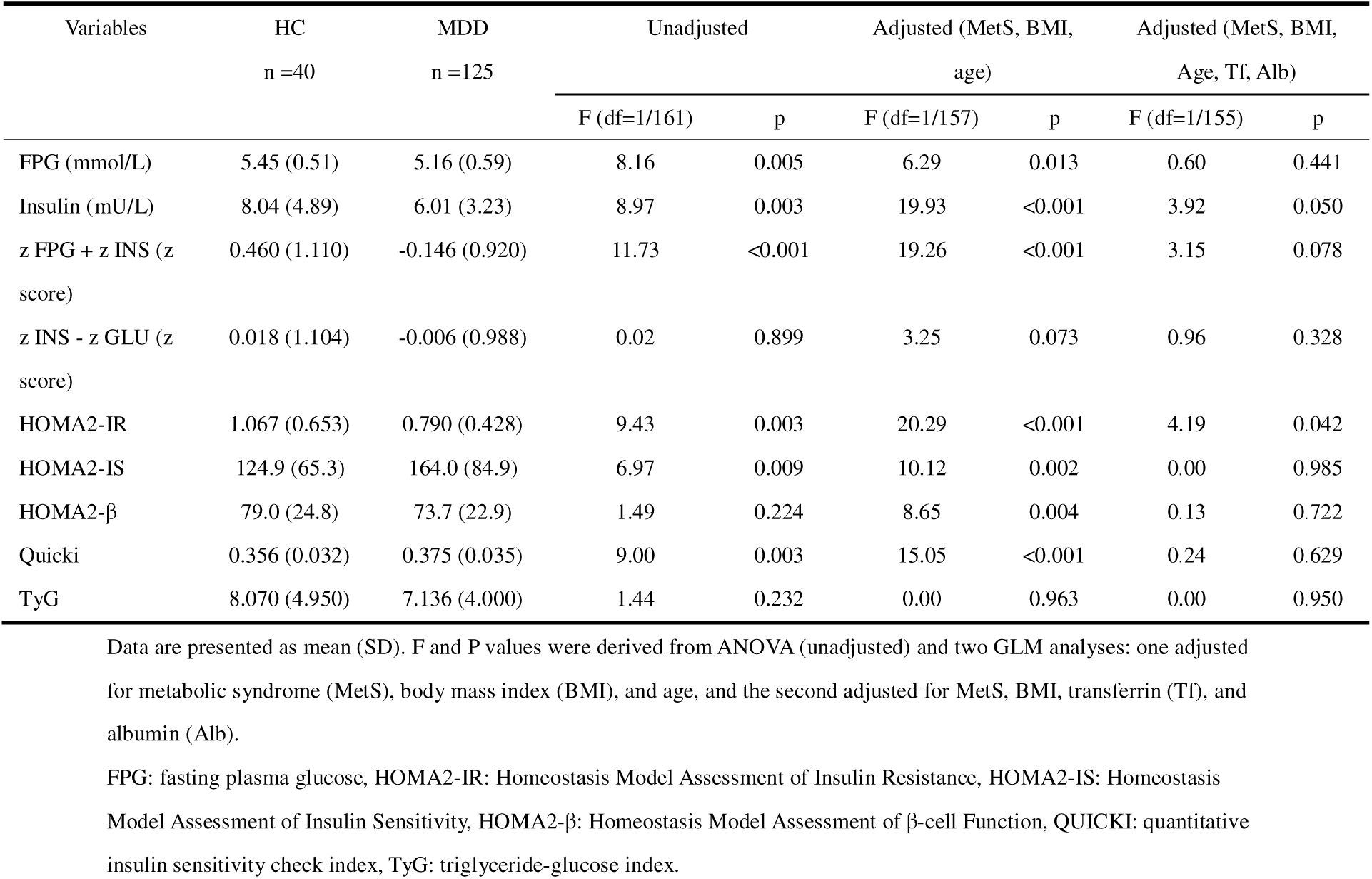
Results of general linear models (GLM) showing unadjusted and adjusted associations between metabolic indicators and major depressive disorder (MDD) versus healthy controls (HC)

| Variables | HC<br>n =40 | MDD<br>n =125 | Unadjusted |  | Adjusted (MetS, BMI,<br>age) |  | Adjusted (MetS, BMI,<br>Age, Tf, Alb) |  |
| --- | --- | --- | --- | --- | --- | --- | --- | --- |
|  |  |  | F (df=1/161) | p | F (df=1/157) | p | F (df=1/155) | p |
| FPG (mmol/L) | 5.45 (0.51) | 5.16 (0.59) | 8.16 | 0.005 | 6.29 | 0.013 | 0.60 | 0.441 |
| Insulin (mU/L) | 8.04 (4.89) | 6.01 (3.23) | 8.97 | 0.003 | 19.93 | <0.001 | 3.92 | 0.050 |
| z FPG + z INS (z<br>score) | 0.460 (1.110) | -0.146 (0.920) | 11.73 | <0.001 | 19.26 | <0.001 | 3.15 | 0.078 |
| z INS - z GLU (z<br>score) | 0.018 (1.104) | -0.006 (0.988) | 0.02 | 0.899 | 3.25 | 0.073 | 0.96 | 0.328 |
| HOMA2-IR | 1.067 (0.653) | 0.790 (0.428) | 9.43 | 0.003 | 20.29 | <0.001 | 4.19 | 0.042 |
| HOMA2-IS | 124.9 (65.3) | 164.0 (84.9) | 6.97 | 0.009 | 10.12 | 0.002 | 0.00 | 0.985 |
| HOMA2-β | 79.0 (24.8) | 73.7 (22.9) | 1.49 | 0.224 | 8.65 | 0.004 | 0.13 | 0.722 |
| Quicki | 0.356 (0.032) | 0.375 (0.035) | 9.00 | 0.003 | 15.05 | <0.001 | 0.24 | 0.629 |
| TyG | 8.070 (4.950) | 7.136 (4.000) | 1.44 | 0.232 | 0.00 | 0.963 | 0.00 | 0.950 |

Data are presented as mean (SD). F and P values were derived from ANOVA (unadjusted) and two GLM analyses: one adjusted for metabolic syndrome (MetS), body mass index (BMI), and age, and the second adjusted for MetS, BMI, transferrin (Tf), and albumin (Alb).
FPG: fasting plasma glucose, HOMA2-IR: Homeostasis Model Assessment of Insulin Resistance, HOMA2-IS: Homeostasis Model Assessment of Insulin Sensitivity, HOMA2-β : Homeostasis Model Assessment of $\beta$ -cell Function, QUICKI: quantitative insulin sensitivity check index, TyG: triglyceride-glucose index.

Our observations indicate that 92 patients received treatment with antidepressants, 10 were treated with mood stabilizers, 68 were administered benzodiazepines, and 44 were prescribed antipsychotic medications. GLM analyses revealed that treatment with these 4 types of drugs did not yield any significant effects on the insulin biomarkers even in the absence of FDR p-value correction. As such, use of these drugs did not affect the biomarker results of the current study.

### Correlations between insulin, negative APPs, metabolic markers, and weight loss

**Table 3** shows the Pearson correlation coefficients among negative APPs, metabolic indicators, anthropometric measures, and weight loss. Plasma levels of transferrin and albumin showed positive correlations with FPG, insulin, z FPG + z insulin, HOMA2-IR, and HOMA2-β. In contrast, both biomarkers were negatively correlated with insulin sensitivity indices (HOMA2-IS and QUICKI). The TyG index was positively correlated with albumin but not with transferrin. The anthropometric measures BMI and WC demonstrated strong positive correlations with insulin resistance parameters and strong negative correlations with insulin sensitivity indices. Weight loss was negatively correlated with FPG, insulin, z FPG + z insulin score, and HOMA2-IR, and positively correlated with QUICKI.

**Table 3.** Intercorrelation matrix (Pearson’s correlation coefficients) between insulin biomarkers, the acute phase response and metabolic indicators.

| Variables | Transferrin | Albumin | BMI | WC | Age | Weight loss |
| --- | --- | --- | --- | --- | --- | --- |
| FPG | 0.192* | 0.243** | 0.458** | 0.425** | 0.366** | -0.225** |
| Insulin | 0.295** | 0.369** | 0.480** | 0.383** | 0.044 | -0.185* |
| z FPG + INS | 0.282** | 0.355** | 0.544** | 0.468** | 0.238** | -0.238** |
| z INS - GLU | 0.102 | 0.125 | 0.021 | -0.042 | -0.317** | 0.039 |
| HOMA2-IR | 0.298** | 0.370** | 0.489** | 0.391** | 0.058 | -0.191* |
| HOMA2-IS | -0.268** | -0.370** | -0.407** | -0.381** | -0.088 | 0.131 |
| HOMA2-β | 0.223** | 0.293** | 0.240** | 0.165* | -0.177* | -0.047 |
| QUICKI | -0.292** | -0.385** | -0.478** | -0.042 | -0.124 | 0.178* |
| TyG | 0.100 | 0.227** | 0.360** | 0.373** | 0.298** | -0.124 |

\*\* Correlation is significant at the 0.01 level (2-tailed); \* Correlation is significant at the 0.05 level (2-tailed).
FPG: fasting plasma glucose, INS: insulin, GLU: glucose, HOMA2-IR: Homeostasis Model Assessment of Insulin Resistance, HOMA2-IS: Homeostasis Model Assessment of Insulin Sensitivity, HOMA2-β : Homeostasis Model Assessment of $\beta$ -cell Function, QUICKI: quantitative insulin sensitivity check index, TyG: triglyceride-glucose index, MetS: Metabolic syndrome.

### Correlations Between Metabolic Indicators and Severity Scales

**Table 4** shows the correlations between metabolic biomarkers and symptom severity scales. FPG and insulin were significantly and negatively correlated with the HAMD, HAMA, FF, and STAI state scales. The z FPG + z insulin and HOMA-2IR scores exhibited significant negative correlations across all these scales. HOMA2-IS was positively correlated with these scales, except HAMA. QUICKI showed significant positive correlations with the severity scales.

**Table 4.**
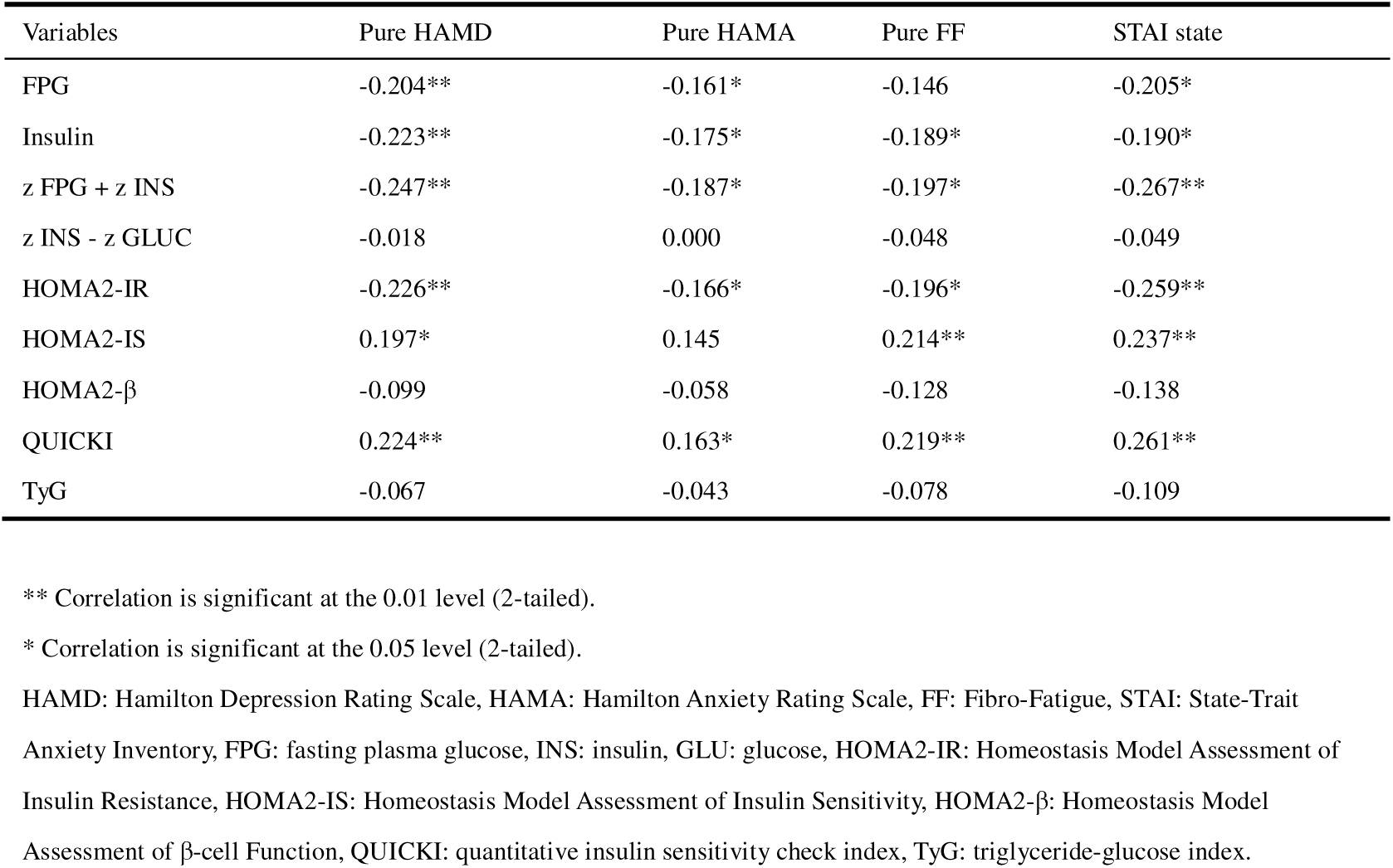
Intercorrelation matrix (Pearson’s correlation coefficients) between biomarkers and severity scales.

| Variables | Pure HAMD | Pure HAMA | Pure FF | STAI state |
| --- | --- | --- | --- | --- |
| FPG | -0.204** | -0.161* | -0.146 | -0.205* |
| Insulin | -0.223** | -0.175* | -0.189* | -0.190* |
| z FPG + z INS | -0.247** | -0.187* | -0.197* | -0.267** |
| z INS - z GLUC | -0.018 | 0.000 | -0.048 | -0.049 |
| HOMA2-IR | -0.226** | -0.166* | -0.196* | -0.259** |
| HOMA2-IS | 0.197* | 0.145 | 0.214** | 0.237** |
| HOMA2-β | -0.099 | -0.058 | -0.128 | -0.138 |
| QUICKI | 0.224** | 0.163* | 0.219** | 0.261** |
| TyG | -0.067 | -0.043 | -0.078 | -0.109 |

\*\* Correlation is significant at the 0.01 level (2-tailed).
\* Correlation is significant at the 0.05 level (2-tailed).
HAMD: Hamilton Depression Rating Scale, HAMA: Hamilton Anxiety Rating Scale, FF: Fibro-Fatigue, STAI: State-Trait Anxiety Inventory, FPG: fasting plasma glucose, INS: insulin, GLU: glucose, HOMA2-IR: Homeostasis Model Assessment of Insulin Resistance, HOMA2-IS: Homeostasis Model Assessment of Insulin Sensitivity, HOMA2-β : Homeostasis Model Assessment of $\beta$ -cell Function, QUICKI: quantitative insulin sensitivity check index, TyG: triglyceride-glucose index.

### Predictors of Metabolic Dysregulation

**Table 5** summarizes the results of multiple regression analyses examining the predictors of metabolic measures. Regression model #1 shows that 35.9% of the variance in FPG was explained by age, transferrin, and WC (all positive associations), together with weight loss (negative association). Regression model #2 demonstrates that 34.8% of the variance in insulin was explained by BMI and albumin (positive associations), along with female gender. Regression model #3 indicates that 36.8% of the variance in HOMA-IR was explained by BMI and albumin (positive associations), together with female gender. Regression model #4 shows that 41.4% of the variance in the z FPG + z insulin score was explained by BMI, albumin, transferrin, and age (all positive associations). Regression model #5 demonstrates that 34.0% of the variance in the QUICKI index was explained by BMI, albumin, and transferrin (all negative associations).

**Table 5.**
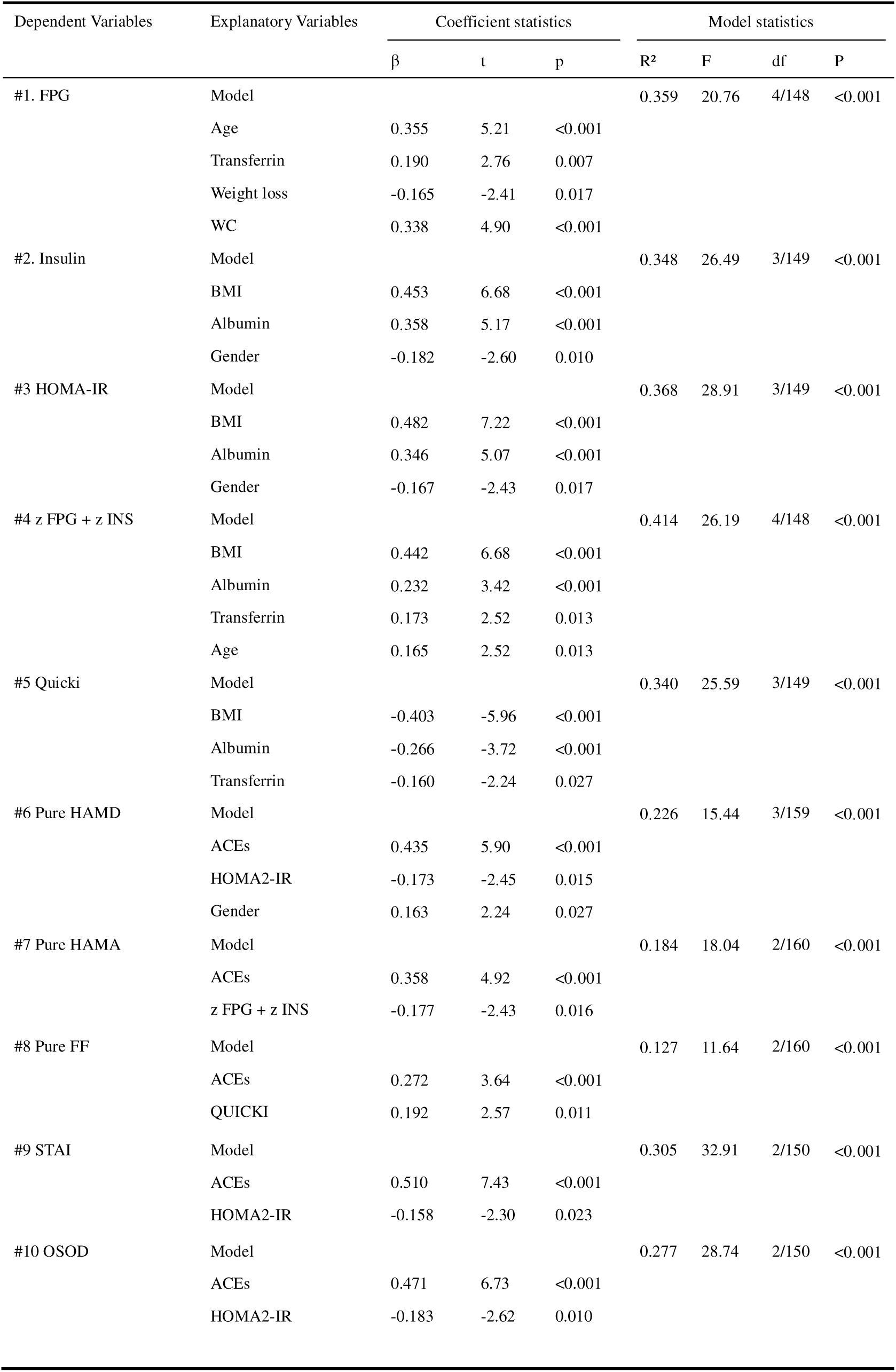
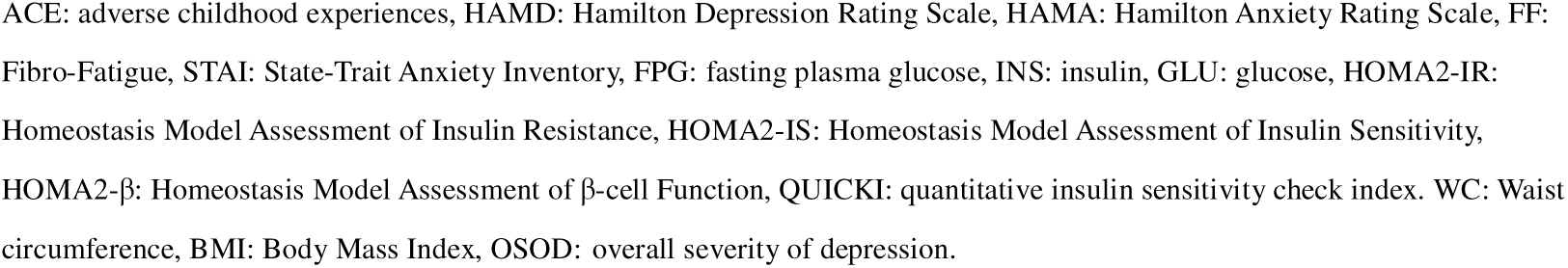
Multiple regression with insulin metabolism biomarkers or rating scale scores as dependent variables.

| Dependent Variables | Explanatory Variables | Coefficient statistics |  |  | Model statistics |  |  |  |
| --- | --- | --- | --- | --- | --- | --- | --- | --- |
| | | $\beta$ | t | p | R<sup>2</sup> | F | df | P |
| #1. FPG | Model |  |  |  | 0.359 | 20.76 | 4/148 | <0.001 |
|  | Age | 0.355 | 5.21 | <0.001 |  |  |  |  |
|  | Transferrin | 0.190 | 2.76 | 0.007 |  |  |  |  |
|  | Weight loss | -0.165 | -2.41 | 0.017 |  |  |  |  |
|  | WC | 0.338 | 4.90 | <0.001 |  |  |  |  |
| #2. Insulin | Model |  |  |  | 0.348 | 26.49 | 3/149 | <0.001 |
|  | BMI | 0.453 | 6.68 | <0.001 |  |  |  |  |
|  | Albumin | 0.358 | 5.17 | <0.001 |  |  |  |  |
|  | Gender | -0.182 | -2.60 | 0.010 |  |  |  |  |
| #3 HOMA-IR | Model |  |  |  | 0.368 | 28.91 | 3/149 | <0.001 |
|  | BMI | 0.482 | 7.22 | <0.001 |  |  |  |  |
|  | Albumin | 0.346 | 5.07 | <0.001 |  |  |  |  |
|  | Gender | -0.167 | -2.43 | 0.017 |  |  |  |  |
| #4 z FPG + z INS | Model |  |  |  | 0.414 | 26.19 | 4/148 | <0.001 |
|  | BMI | 0.442 | 6.68 | <0.001 |  |  |  |  |
|  | Albumin | 0.232 | 3.42 | <0.001 |  |  |  |  |
|  | Transferrin | 0.173 | 2.52 | 0.013 |  |  |  |  |
|  | Age | 0.165 | 2.52 | 0.013 |  |  |  |  |
| #5 Quicki | Model |  |  |  | 0.340 | 25.59 | 3/149 | <0.001 |
|  | BMI | -0.403 | -5.96 | <0.001 |  |  |  |  |
|  | Albumin | -0.266 | -3.72 | <0.001 |  |  |  |  |
|  | Transferrin | -0.160 | -2.24 | 0.027 |  |  |  |  |
| #6 Pure HAMD | Model |  |  |  | 0.226 | 15.44 | 3/159 | <0.001 |
|  | ACEs | 0.435 | 5.90 | <0.001 |  |  |  |  |
|  | HOMA2-IR | -0.173 | -2.45 | 0.015 |  |  |  |  |
|  | Gender | 0.163 | 2.24 | 0.027 |  |  |  |  |
| #7 Pure HAMA | Model |  |  |  | 0.184 | 18.04 | 2/160 | <0.001 |
|  | ACEs | 0.358 | 4.92 | <0.001 |  |  |  |  |
|  | z FPG + z INS | -0.177 | -2.43 | 0.016 |  |  |  |  |
| #8 Pure FF | Model |  |  |  | 0.127 | 11.64 | 2/160 | <0.001 |
|  | ACEs | 0.272 | 3.64 | <0.001 |  |  |  |  |
|  | QUICKI | 0.192 | 2.57 | 0.011 |  |  |  |  |
| #9 STAI | Model |  |  |  | 0.305 | 32.91 | 2/150 | <0.001 |
|  | ACEs | 0.510 | 7.43 | <0.001 |  |  |  |  |
|  | HOMA2-IR | -0.158 | -2.30 | 0.023 |  |  |  |  |
| #10 OSOD | Model |  |  |  | 0.277 | 28.74 | 2/150 | <0.001 |
|  | ACEs | 0.471 | 6.73 | <0.001 |  |  |  |  |
|  | HOMA2-IR | -0.183 | -2.62 | 0.010 |  |  |  |  |

ACE: adverse childhood experiences, HAMD: Hamilton Depression Rating Scale, HAMA: Hamilton Anxiety Rating Scale, FF: Fibro-Fatigue, STAI: State-Trait Anxiety Inventory, FPG: fasting plasma glucose, INS: insulin, GLU: glucose, HOMA2-IR: Homeostasis Model Assessment of Insulin Resistance, HOMA2-IS: Homeostasis Model Assessment of Insulin Sensitivity, HOMA2-β : Homeostasis Model Assessment of $\beta$ -cell Function, QUICKI: quantitative insulin sensitivity check index. WC: Waist circumference, BMI: Body Mass Index, OSOD: overall severity of depression.

### Predictors of the clinical rating scale scores

Table 5, regression model #6 indicates that 22.6% of the variance in the pure HAMD score was explained by ACEs and female gender (positive associations), together with HOMA-IR (negative association). Regression #7 shows that 18.4% of the variance in the pure HAMA score was explained by ACEs (positive association) and the z FPG + z insulin score (negative association). Model #8 demonstrates that 12.7% of the variance in the pure FF score was explained by ACEs and the QUICKI index (both positive associations). Model #9 indicates that 30.5% of the variance in the STAI score was explained by ACEs (positive association) and HOMA2-IR (negative association). Regression #10 shows that 27.7% of the variance in the OSOD score was explained by ACEs (positive association) and HOMA2-IR (negative association).

### Impact of metabolic syndrome on insulin biomarkers in MDD

**Electronic Supplementary File** (ESF), Table 1 shows the findings from general linear models assessing the impact of MetS on insulin biomarkers, after controlling for potential confounders including MDD. Individuals with MetS showed significantly higher FPG, insulin, z FPG + z insulin, HOMA2-IR, and TYG than those without MetS, whilst HOMA2S and the QUICKI index were significantly lower in those with MetS.

## Discussion

### Insulin resistance and sensitivity in Chinese MDD patients

The first major finding of this study is that Chinese MDD patients exhibit lower insulin resistance than controls, whilst their insulin sensitivity was higher. Whether unadjusted or adjusted for confounding factors such as age, gender, BMI, and MetS, the MDD group showed significantly lower FPG, insulin levels, and HOMA2-IR scores compared to the healthy controls. Meanwhile insulin sensitivity indices, such as HOMA2-IS and QUICKI, were significantly higher.

In contrast, a meta-analysis by Fernandes et al. (Fernandes et al., 2022) reported that insulin levels, HOMA-IR, and insulin variability were significantly elevated in the acute phase of depression. However, this meta-analysis study did not adjust for potential confounding factors such as MetS or increased BMI scores (Fernandes et al., 2022). It is obvious that the absence of correction for metabolic factors might result in significant heterogeneity and bias. It is well established that these factors largely affect insulin resistance and sensitivity and that after controlling for these variables no significant differences between MDD (and BD) and controls may be established (de Melo et al., 2017; Landucci Bonifácio et al., 2017; Morelli et al., 2021). Nevertheless, after considering the effects of other NIMETOX pathways (e.g., immune, and oxidative stress), it was established that insulin resistance is associated with the severity of illness (Maes, Jirakran, et al., 2025b). Therefore, the results of most studies included in the meta-analysis by Fernandes et al. are difficult to interpret. After adjusting for BMI, MetS and age (sex was not significant) the current study found that the increase in insulin sensitivity and lower insulin resistance were still prevalent in our Chinese MDD patients.

The above-mentioned discrepancies may be due to significant differences in obesity and MetS prevalence between our study sample and other samples recruited from Western or some South-American countries. Epidemiological data show that the age-standardized prevalence of obesity in China (approximately 16.0% in men and 14.4% in women) (Mu et al., 2021) is significantly lower than in Western countries (e.g., approximately 43.0% in men and 42.1% in women in the United States) (Hales et al., 2020). Furthermore, in Chengdu, China the prevalence of obesity is approximately 11.0% and 16.7% for MetS (Zhou et al., 2024) significantly lower than the 37.6% seen in U.S (Liang et al., 2023). Our representative cohort from Chengdu exhibited a comparatively low prevalence of MetS, comprising 17.7% of MDD patients and 25.6% of the controls. The lower incidence of obesity and MetS might in theory maybe explain the observed differences in insulin sensitivity in Chinese MDD patients versus studies that did not adjust for MetS or obesity. Nonetheless, the exact explanation is more intricate, as detailed in the subsequent two sections.

### Insulin sensitivity and the APP response

The second major finding of this study is the significant association between enhanced insulin resistance in MDD and the negative APP. Despite the lower FPG, insulin levels, and HOMA2-IR scores in MDD, and the significantly increased insulin sensitivity, the significance of these variables disappeared after adjusting for negative APPs. This indicates that the APP response impacts insulin metabolism and might explain the lower insulin resistance and higher insulin sensitivity in MDD.

Several studies have reported a decrease in albumin and transferrin levels in depression (Al-Marwani et al., 2023; Ambrus & Westling, 2019; Maes, 1993; Maes et al., 1991; Van Hunsel et al., 1996; Yin et al., 2022). Maes et al. (1991) found that depression is associated with a negative APP response and protein deficiency or homeostasis dysregulation, which additionally was associated with anorexia and weight loss, core symptoms of depression (Maes et al., 1991). Our study also supports this hypothesis, suggesting that protein malnutrition associated with weight loss is associated with the negative APP response in MDD.

Additionally, recent research indicates that the reduction in negative APPs in MDD patients, along with changes in positive APPs (such as high-sensitivity monomeric C-reactive protein), reflects a mild smoldering inflammatory state (Maes, Niu, et al., 2025c). This state may be associated with chronic malnutrition, the activation of chronic inflammatory factors (such as TNF-α and IL-6), and impaired liver synthesis function of positive APPs (Cao et al., 2022; Maes, Almulla, et al., 2025a; Maes et al., 1991). The liver is the primary organ responsible for synthesizing negative APPs (Ceciliani et al., 2002). Under normal physiological conditions, the liver efficiently synthesizes and secretes these proteins to maintain metabolic balance and immune function. However, in MDD, chronic inflammatory factors may inhibit liver function, resulting in a reduction in the synthesis of negative APPs (Maes, 1993; Maes et al., 1991; Maes, Niu, et al., 2025c; Shao et al., 2021). For example, TNF-α can suppress the synthesis of albumin and transferrin by activating the Nuclear Factor-κB signaling pathway, resulting in significantly lower plasma concentrations of these proteins (Chojkier, 2005).

This inflammation-mediated inhibition of liver synthesis may lead to a “malnutrition-inflammation” vicious cycle. Albumin and transferrin are not only important nutritional markers but also play a key function in sustaining homeostasis and various physiological functions (Levitt & Levitt, 2016; Talukder, 2021). Their sustained low levels may exacerbate the pathogenesis of MDD. On one hand, hypoalbuminemia may lead to an imbalance in colloid osmotic pressure, reduced oxidative stress buffering capacity, and impaired toxin clearance (Dubois et al., 2006; Levitt & Levitt, 2016). On the other hand, reduced transferrin may impair iron transport and metabolism, increasing the risk of the anemia of inflammation (Maes et al., 1996) and affecting mitochondrial energy production, further exacerbating fatigue in MDD patients (Rybka et al., 2013; Talukder, 2021).

### Metabolic flexibility and enhanced insulin sensitivity

Metabolic flexibility refers to the ability of the organism to maintain normal physiological function by adjusting energy metabolism in the context of chronic stress or chronic disease (Smith et al., 2018). This adaptive mechanism involves the reallocation of energy resources to prioritize the needs of critical organs (Smith et al., 2018). Particularly under the stress of chronic inflammation and protein malnutrition, the body may optimize the use of limited energy resources by altering metabolic pathways, especially to ensure the energy supply of vital organs such as the brain (Olson et al., 2020).

In this study, we observed abnormally elevated insulin sensitivity in MDD in the context of a negative APP response, suggesting that this change in metabolic status may be a metabolic adaptive response of the body. Long-term activation of chronic inflammatory factors may have metabolic consequences, leading to adaptive adjustments in energy allocation and use by the organism (Straub, 2011; Wang & Ye, 2015). In response to a chronic inflammatory state, the organism may optimize glucose utilization by increasing insulin sensitivity and reducing the waste of energy resources (Berbudi et al., 2025).

In people without MetS, mild, smoldering inflammation can transiently lower insulin resistance. For example, small elevations in IL-6 may activate AMPK and increase insulin-stimulated glucose disposal in humans, improving glucose uptake (Carey et al., 2006). This hormetic window closes as inflammation strengthens or when MetS is present (Koenen et al., 2021). In MetS, the same stimulus typically worsens insulin resistance because inflammatory pathways are pre-primed, including JNK/IKKβ signaling, macrophage infiltration, and lipotoxic mediators blunting PI3K-Akt thereby promoting hepatic and muscle insulin resistance (Kelly et al., 2009; Pedersen & Febbraio, 2008; Shoelson et al., 2006). In adipose tissue, proinflammatory remodeling with macrophage infiltration elevates serum cytokines and free fatty acids, reinforcing systemic insulin resistance dysfunction (Apovian et al., 2008). Furthermore, reciprocal interactions between neurotoxicity-associated cytokine networks and MetS are associated with increasing clinical severity of MDD (Maes, Jirakran, et al., 2025b). Overall, mild smoldering inflammation may be hormetic with respect to insulin metabolism in subjects without MetS, but in those with MetS it might, via interactions among activated cytokine networks and MetS, sustain or aggravate insulin resistance and severity of MDD (Maes, Jirakran, et al., 2025b).

#### Limitations

This cross-sectional study has several limitations. First, this is a case-control study which limits the ability to draw definitive conclusions about causality. Moreover, this study was conducted at a single site in southwestern China, and lifestyle factors (such as dietary intake, dietary habits, and physical activity) in this region not only differ from the West but may also differ from those in other Chinese areas.

## Conclusions

This study shows that Chinese MDD patients exhibit enhanced insulin sensitivity and lower insulin resistance. Furthermore, these changes are mediated by a mild smoldering inflammatory response with protein malnutrition. A mild inflammatory response in study samples without a high prevalence of subjects with MetS or obesity (as in our Chengdu study sample) may be accompanied by a hormetic response briefly enhancing insulin sensitivity. On the other hand, stronger long-standing inflammatory responses, especially in populations with increased prevalence of MetS and obesity (such as Western populations) might cause and maintain impaired insulin resistance and cause increased severity of depression.

Therefore, under the dual stress of chronic inflammation and protein malnutrition, MDD is characterized by increased insulin efficiency by prioritizing energy supply to critical organs, thereby optimizing glucose resource utilization to maintain normal function of vital organs, especially when energy supply is limited. However, the presence of MetS or obesity obscures these primary biomarkers of MDD, since interactions between immunological and metabolic factors become more pronounced. Further research should examine these immune and MetS interactions in MDD.

Consequently, it is likely erroneous to assert that MDD is inherently linked to heightened insulin resistance; rather, the accurate interpretation is that MDD in the absence of MetS is accompanied by increased insulin sensitivity, and in the presence of immune activation and MetS might elicit elevated insulin resistance, which affects the severity of the condition.

## Supporting information

Electronic Supplementary File

## Data Availability

The dataset supporting this study is available from the corresponding author (MM) upon reasonable request and after a thorough data review.

## Acknowledgements

Not Applicable

## Author’s contributions

Each author made an equal contribution to this study and has reviewed and agreed to the submitted version of the manuscript.

