## Supplementary material for "Lower insulin resistance in Chinese patients with severe major depressive disorder: associations with the inflammatory response": Electronic Supplementary File

**Electronic Supplementary File (ESF)**

ESF, Table 1. Differences in insulin biomarkers between subjects with and without metabolic syndrome (MetS).

| Variables | MetS | MetS | F | df | P |
| --- | --- | --- | --- | --- | --- |
| FPG (mmol/L) | 5.748 (0.094) | 5.167 (0.047) | 28.65 | 1/157 | < 0.001 |
| Insulin (mU/L) | 9.691 (0.655) | 6.064 (0.327) | 23.16 | 1/157 | < 0.001 |
| z FPG + z INS | 1.016 (0.156) | -0.124 (0.078) | 40.14 | 1/157 | < 0.001 |
| z INS - z GLU | -0.059 (0.202) | -0.012 (0.101) | 0.04 | 1/157 | 0.842 |
| HOMA2-IR | 1.299 (0.086) | 0.797 (0.043) | 25.67 | 1/157 | < 0.001 |
| HOMA2-IS | 104.944 (15.550) | 157.242 (7.760) | 8.55 | 1/157 | 0.004 |
| HOMA2-β | 81.507 (4.716) | 74.047 (2.353) | 1.89 | 1/157 | 0.171 |
| Quicki | 0.344 (0.006) | 0.373 (0.003) | 16.59 | 1/157 | < 0.001 |
| TyG | 13.047 (0.666) | 6.040 (0.332) | 83.75 | 1/157 | < 0.001 |

Data are presented as marginal estimated mean (SE); F and P values were derived from GLM adjusted for body mass index and age, without MetS; df: degrees of freedom are reported for each F-test.

FPG: fasting plasma glucose, INS: insulin, GLU: glucose, HOMA2-IR: Homeostasis Model Assessment of Insulin Resistance, HOMA2-IS: Homeostasis Model Assessment of Insulin Sensitivity, HOMA2-β: Homeostasis Model Assessment of β-cell Function, QUICKI: quantitative insulin sensitivity check index, TyG: triglyceride-glucose index, MetS: Metabolic syndrome.
